# Machine learning analysis highlights the down-trending of the proportion of COVID-19 patients with a distinct laboratory result profile

**DOI:** 10.1101/2020.11.28.20240150

**Authors:** He S. Yang, Yu Hou, Hao Zhang, Amy Chadburn, Lars F. Westblade, Richard Fedeli, Peter A.D. Steel, Sabrina E. Racine-Brzostek, Priya Velu, Jorge L. Sepulveda, Michael J. Satlin, Melissa M. Cushing, Rainu Kaushal, Zhen Zhao, Fei Wang

**Affiliations:** Department of Pathology and Laboratory Medicine, Weill Cornell Medicine, New York, NY, USA; New York-Presbyterian Hospital/Weill Cornell Medical Campus, New York, NY, US; Department of Population Health Sciences, Weill Cornell Medicine, New York, NY, USA; Department of Medicine, Weill Cornell Medicine, New York, NY, US; Department of Emergency Medicine, Weill Cornell Medicine, New York, NY, USA; Deparment of Pathology and Laboratory Informatics, School of Medicine and Health Sciences, George Washington University, Washington DC, US; Division of Infectious Disease, Department of Medicine, Weill Cornell Medicine, New York, NY, US

**Keywords:** Coronavirus Disease 2019 (COVID-19), machine learning, Unified Manifold Approximation and Project (UMAP), routine laboratory tests, severe acute respiratory syndrome coronavirus 2 (SARS-CoV-2)

## Abstract

**Background:** New York City (NYC) experienced an initial surge and gradual decline in the number of SARS-CoV-2 confirmed cases in 2020. A change in the pattern of laboratory test results in COVID-19 patients over this time has not been reported or correlated with patient outcome.

**Methods:** We performed a retrospective study of routine laboratory and SARS-CoV-2 RT-PCR test results from 5,785 patients evaluated in a NYC hospital emergency department from March to June employing machine learning analysis.

**Results:** A COVID-19 high-risk laboratory test result profile (COVID19-HRP), consisting of 21 routine blood tests, was identified to characterize the SARS-CoV-2 patients. Approximately half of the SARS-CoV-2 positive patients had the distinct COVID19-HRP that separated them from SARS-CoV-2 negative patients. SARS-CoV-2 patients with the COVID19-HRP had higher SARS-CoV-2 viral loads, determined by cycle-threshold values from the RT-PCR, and poorer clinical outcome compared to other positive patients without COVID19-HRP. Furthermore, the percentage of SARS-CoV-2 patients with the COVID19-HRP has significantly decreased from March/April to May/June. Notably, viral load in the SARS-CoV-2 patients declined and their laboratory profile became less distinguishable from SARS-CoV-2 negative patients in the later phase.

**Conclusions:** Our study visualized the down-trending of the proportion of SARS-CoV-2 patients with the distinct COVID19-HRP. This analysis could become an important tool in COVID-19 population disease severity tracking and prediction. In addition, this analysis may play an important role in prioritizing high-risk patients, assisting in patient triaging and optimizing the usage of resources.

## Introduction

The coronavirus disease-2019 (COVID-19), caused by the severe acute respiratory syndrome coronavirus 2 (SARS-CoV-2) (1), has rapidly spread across the globe resulting in 40.8 million confirmed cases and 1.1 million total deaths as of October 21, 2020 (2). The United States has more confirmed cases than any other country worldwide. New York, which was the initial epicenter of the COVID-19 pandemic and has reported the highest number of death in the U.S. (3), has experienced a gradual decline in the number of cases in the months following the initial surge (4, 5). It is unclear if the decline in total Emergency Department (ED) visits for COVID-19-like illnesses (6) and COVID-19-associated hospitalizations (7) is related to changes in virus virulence, early preferential infection of more vulnerable populations, effectiveness of containment measures, or treatment changes. However, there have been only limited studies describing trends in objective clinical data in COVID-19 patients corresponding to these epidemiologic changes.

Currently in most hospital EDs, patients with symptoms suspicious for COVID-19 undergo a SARS-CoV-2 reverse transcription-polymerase chain reaction (RT-PCR) test and a panel of routine laboratory tests. While the pathophysiology of this new virus is still poorly understood, some of its effects on the human body are reflected in abnormal laboratory values. Several studies (8-10) have reported a number of abnormal routine laboratory test results in SARS-CoV-2 infected patients upon initial evaluation, including changes in the complete blood count (CBC), an increase in inflammatory markers and alterations in albumin and globulin levels. Whether the laboratory characteristics of SARS-CoV-2 infected patients have also shifted with the epidemiological changes over time, reflecting the evolution of COVID-19, remains unknown.

Machine learning algorithms have been successfully utilized in healthcare (11-13) and are powerful applications for predicting SARS-CoV-2 infection status (10, 14), disease progression and mortality (15). They are particularly useful in identifying hidden relationships based on complex sets of variables. As routine laboratory test results provide objective and quantifiable chacterization of the effects of the virus on the human body, our study aimed to elucidate the trending of COVID-19 from a laboratory testing prospective. Using machine learning analysis, we identified a distinct panel of abnormal test results (COVID-19 typical laboratory test result profile; COVID19-TLP), which separate SARS-CoV-2 positive from SARS-CoV-2 negative patients and visualized the temporal changes in the COVID19-TLP of SARS-CoV-2 positive patients from the initial outbreak in March and April to a post-apex phase in May and June 2020.

## Methods

### Ethics Statement

This study was approved by the Weill Cornell Medicine Institutional Review Board (#20-03021671).

### Patient cohort and data collection

The test results analyzed in this study were from 5,785 patients evaluated in the ED of New York Presbyterian Hospital/Weill Cornell Medical Center (NYPH/WCMC) from March 11 to June 30, 2020. SARS-CoV-2 RT-PCR results, routine laboratory testing results, patient demographic information (age, sex and race, Table 1), and clinical outcome (hospital admission, ICU admission, mechanical intubation, survival/death) were obtained from the laboratory information system (Cerner Millennium, Cerner Corporation, North Kansas City, Missouri, US). Since the turn-around time (TAT) of RT-PCR is up to 24 hours in our institution whereas the results of routine laboratory testing are usually available within 1-2 hours, laboratory testing results performed within a 48-hour window (± 24 hours) of completion of each RT-PCR test were used in the data analysis. Exclusion criteria included patients < 18 years old, patients who had indeterminate RT-PCR results [RT-PCR positive for the pan-Sarbecovirus target (E gene), yet negative for the SARS-CoV-2 specific target], and patients who did not have any laboratory test results within the time frame (inclusion/exclusion cascade, **Figure 1**). In total, our dataset included the routine laboratory test results from 1,309 SARS-CoV-2 RT-PCR positive and 3,658 RT-PCR negative patients (total 4,967 patients) who ranged in age from 18 to 104 years (median = 60.0 years). Violin plots of the age distribution in all patients as well as SARS-CoV-2 positive patients during the 4 study months are shown in **Supplemental Figure 1**.

**Table 1.**
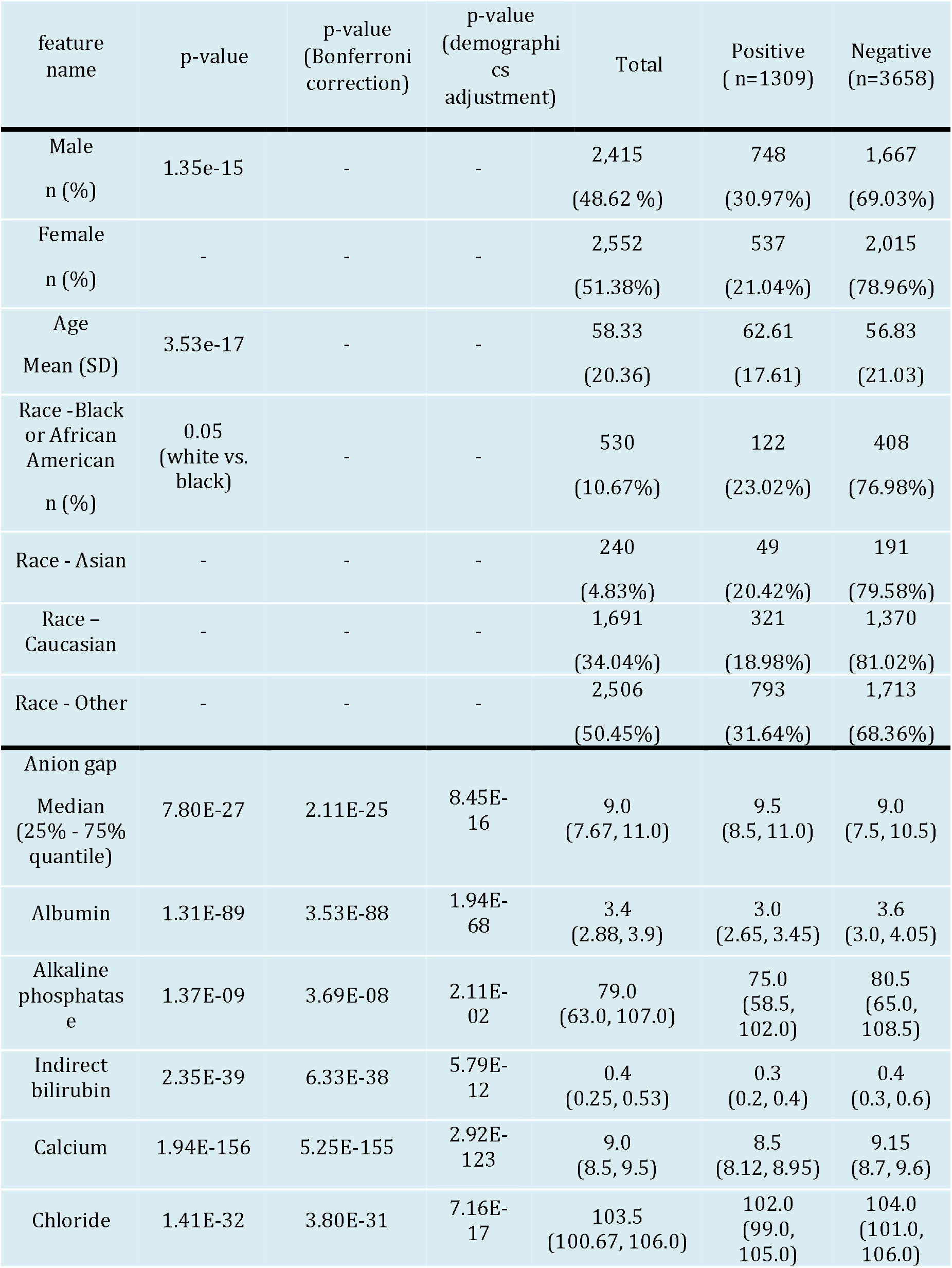
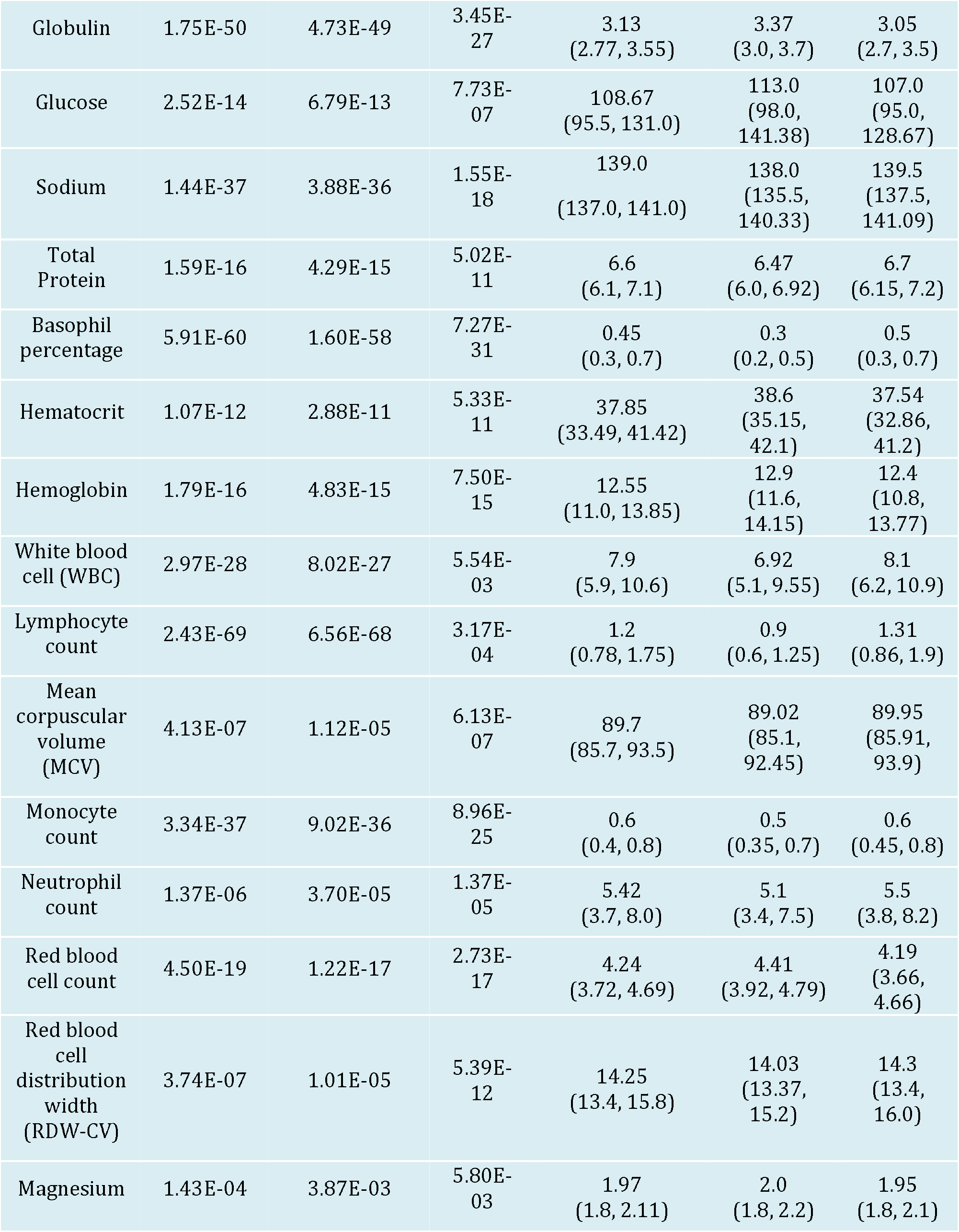
Demographic information of the patient cohort and comparison of 21 laboratory tests in SARS-CoV-2 positive and negative patients.

**Figure 1.**
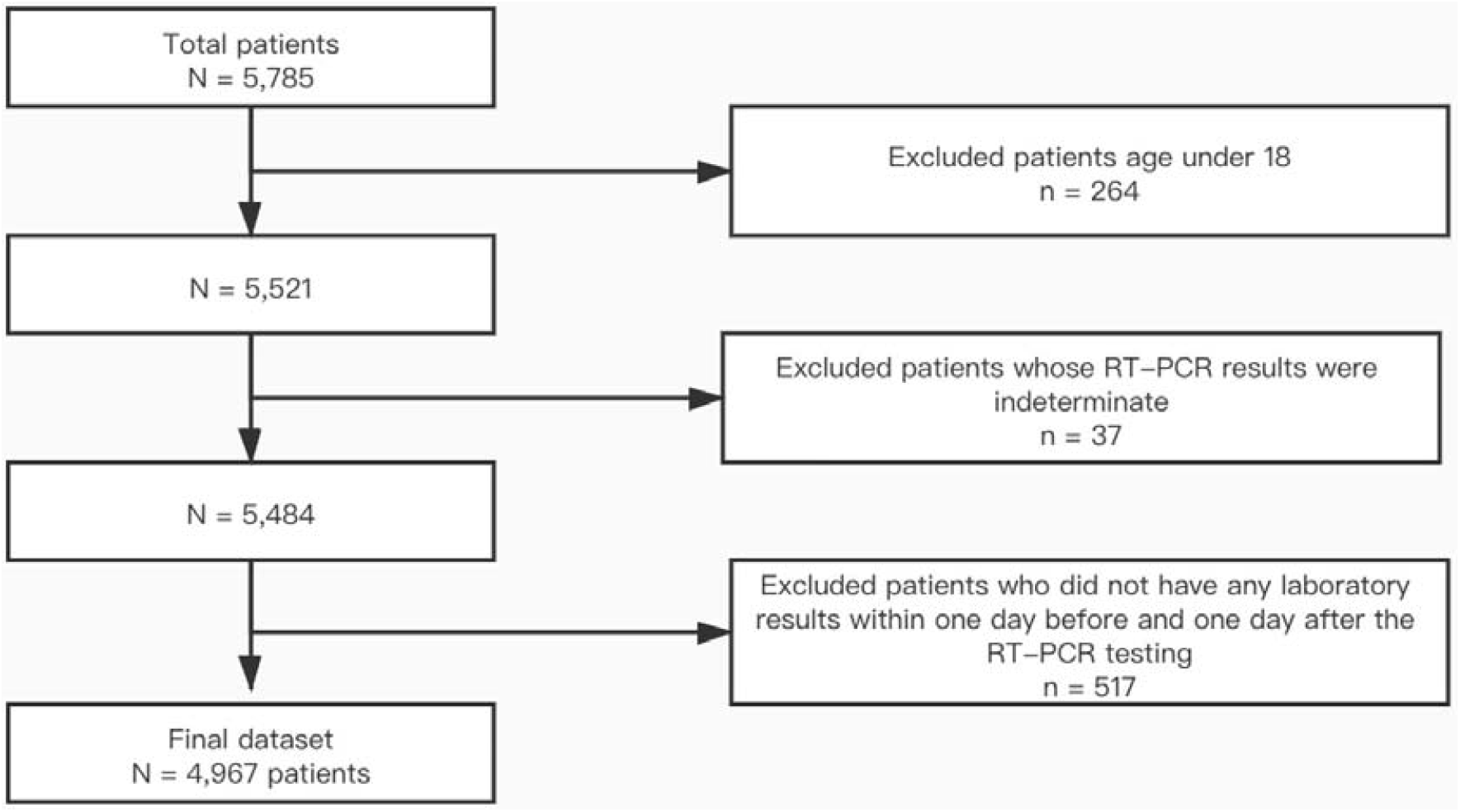
Inclusion/exclusion cascade of patients in the dataset.

### SARS-CoV-2 RT-PCR testing

SARS-CoV-2 RT-PCR testing was performed at NYPH/WCMC using the RealStar SARS-CoV-2 RT-PCR Kit 1.0 reagent system (Altona, Hamburg, Germany) which targets on the S gene and E gene, the Cobas SARS-CoV-2 Assay (Roche Molecular Systems, Inc., Branchburg, NJ) which targets the ORF1ab and E genes, and the Xpert Xpress SARS-CoV-2 Assay (Cepheid, Inc., Sunnyvale, CA) which targets the N2 and E genes (16). The ORF1ab and N2 genes are specific for SARS-COV-2, while the E gene is a pan-Sarbecovirus marker. Based upon previous data(16), the diagnostic performance of both the Cobas 6800 and the Xpert Xpress SARS-CoV-2 assays are considered equivalent. SARS-CoV-2 RT-PCR cycle threshold (C_*T*_) values of the SARS-CoV-2 specific target, which correlate inversely with the quantitative viral load (17), were obtained using the cobas SARS-CoV-2 Assay and Xpert Xpress SARS-CoV-2 RT-PCR Assay, as the values for the SARS-CoV-2 specific gene were comparable between platforms (16). C_*T*_ values from the RealStar SARS-CoV-2 RT-PCR assay were excluded from the analysis as the values are not directly comparable to the other two platforms.

### Routine laboratory testing

Routine chemistry testing was performed on the Siemens ADVIA XPT and Centaur XP analyzers (Siemens Healthineers Global, Erlangen, Germany). Procalcitonin was performed on the Roche e411 analyzer (Roche Diagnostics, Indianapolis, IN). Blood gas analysis was performed on the GEM Premier 4000 analyzer (Instrumentation Laboratory, Bedford, MA). Routine hematology testing was performed on the UniCel DXH 800 analyzer (Beckman Coulter, Brea, CA). Coagulation tests were performed on the Instrumentation Laboratory ACLTM TOP CTS Coagulation System.

### The the Unified Manifold Approximation and Projection (UMAP) analysis

Twenty-one laboratory tests were selected from a total of 685 tests that were ordered for all patients in the dataset based on the following criteria: 1) the test result was available for at least 70% of the patients within 48 hours prior to or after a specific SARS-CoV-2 RT-PCR test in each month, and 2) the test result was significantly different (i.e., P-value, P-value after Bonferroni correction, or P-value after demographics adjustment less than 0.05) in patients with a positive SARS-CoV-2 RT-PCR study compared to persons who had a negative result (**Table 1**). If one specific test was ordered multiple times within 48 hours, an average of the values was calculated and used for analysis. The missing value of a specific laboratory test in a feature vector was imputed by the median value of the available non-missing values of that dimension over all patients. Finally, a 21-dimensional vector was constructed to represent every SARS-CoV-2 RT-PCR testing result, which is a unique laboratory test result profile that characterizes each patient.

We then mapped the vectors of all RT-PCR tests onto a two-dimensional space using the UMAP approach (18), with the goal of visualizing the geometric distributions of the RT-PCR test profiles. These profiles were first standardized with z-score scaling (19) before being incorporated into the UMAP algorithm to eliminate the value range discrepancies among different routine laboratory tests. The UMAP analysis allows the geometric topology among the high dimensional vectors to remain in the low dimensional space so that the geometric relationships among the sample vectors can be visually inspected. Therefore, RT-PCR results with similar routine laboratory profiles remain nearby in the embedding space whereas those with distinct laboratory profiles are located at a distance.

After all RT-PCR profiles were projected onto the two-dimensional space, we used Density-Based Spatial Clustering of Applications with Noise (DBSCAN) (20) to identify the high-density region of positive tests. Then we fitted a two-dimensional Gaussian distribution to define a circle in the two-dimensional embedding space. The mean vector and covariance matrix of this Gaussian distribution is [7.01, 4.76] and [(0.55 0.06), (0.06 0.41)], respectively. After having the Gaussian distribution, we plotted its contour lines for probability density function (pdf). Starting from the contour line with the largest pdf value (0.33), which is the mean point, we gradually expanded the contour line with a decrease of the pdf value in a step size of 0.01. In this expanding process, if we found that the number of negative tests was larger than that of positive ones, we would stop and regard this contour line as the circle.

### Statistical analysis

Comparison of the percentages of RT-PCR results within versus outside the circle in each month was performed by Fisher’s exact test and posthoc analysis. Comparison of the C_*T*_ values and length of hospital stay within versus outside the circle was performed by t-test. Comparison of the percentage of SARS-CoV-2 positive patients with or without the circle for hospital admission from ED, percentage of patients required for care in the ICU and mechanical intubation were performed by the Fisher’s exact test, where the p values were obtained after age adjustment. Statistical analysis was performed using Python version 3.7.

## Results

A retrospective analysis of laboratory tests was performed in a final dataset of 1,309 SARS-CoV-2 RT-PCR confirmed positive patients and 3,658 negative patients (**Figure 1**). A summary of the 21 laboratory tests used to construct the 21-dimensional vector representing the COVID19-TLP is shown in **Table 1**. Using the UMAP analysis, we then mapped the vectors of 5,588 RT-PCR tests onto a two-dimensional space. As shown in **Figure 2**, 45% (n = 513) of the overall SARS-CoV-2 RT-PCR positive results clustered in the area within the black circle which depicts the high density region of positive RT-PCR results. The patients who had positive RT-PCR results within the circle showed a distinct laboratory test result profile (COVID19-TLP) different from those individuals with negative RT-PCR results. In contrast, only 3% (n = 116) of SARS-CoV-2 negative RT-PCR results shared the COVID19-TLP and were within the circle. We further performed the UMAP analysis for each of the four months (March, April, May and June) and observed a dramatic change over time: approximately half of the RT-PCR positive results in March (51%) and April (52%) clustered within the circle. When transitioning into May, while the total number of positive cases was declining, positive RT-PCR results associated with COVID19-TLP became significantly fewer, with only 16% of positive RT-PCR results in the circle (p < 0.001 compared to March or April, respectively). In June, the percentage of SARS-CoV-2 RT-PCR positive results in the circle was even less (5%, p = 0.03 compared to May) and relatively more positive RT-PCR were indistinguishably intermixed with the negative RT-PCR results based on the laboratory test result profile. However, it is important to note that more than 90% of the SARS-CoV-2 RT-PCR negative results (97% overall, 92% in March, 96% in April, 98% in May, and 97% in June) fell outside the circle throughout the initial and subsequent months of the SARS-CoV-2 pandemic.

**Figure 2.**
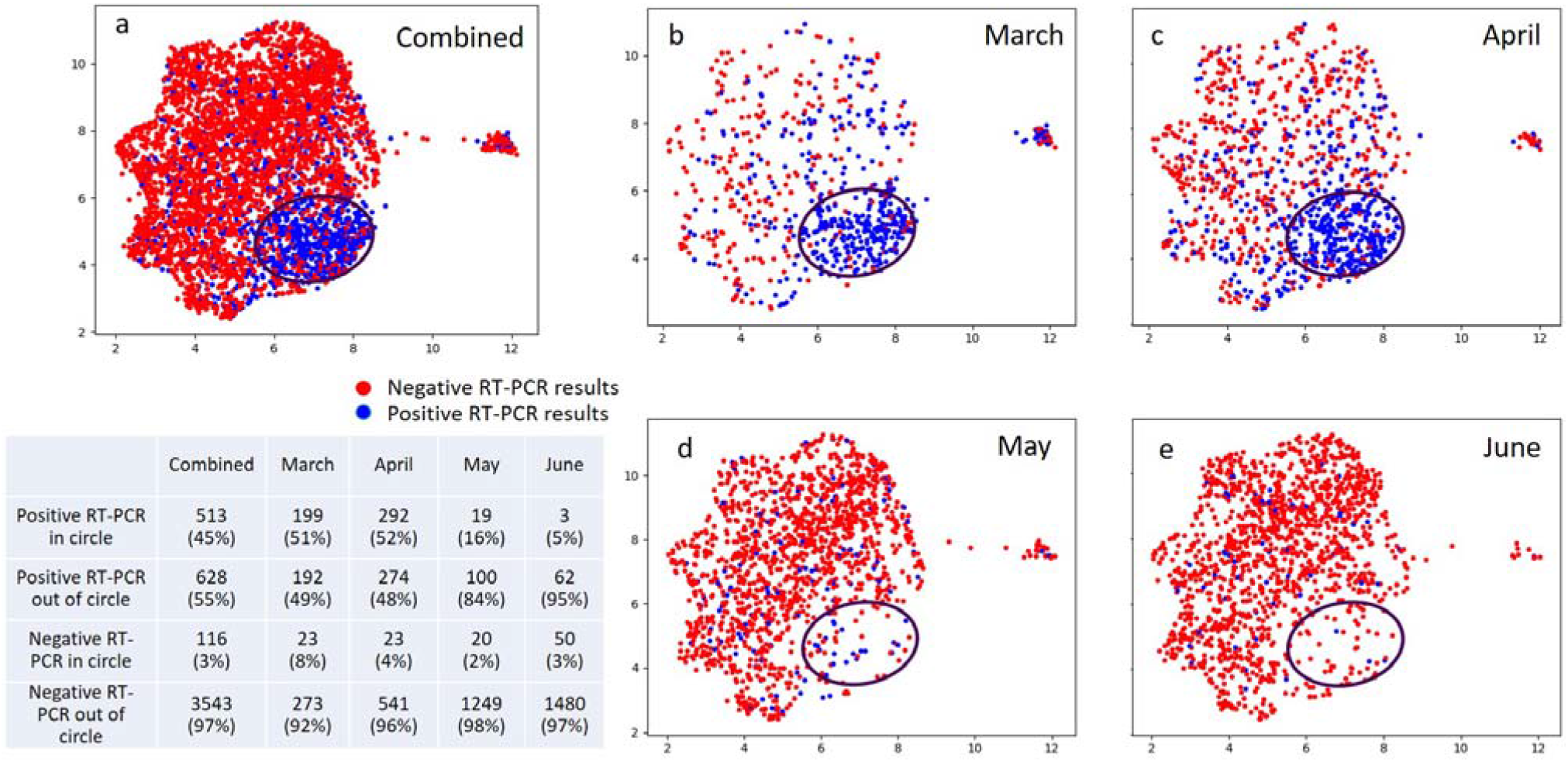
Unified Manifold Approximation and Projection (UMAP) analysis of the laboratory profiles associated with the SARS-CoV-2 RT-PCR positive and negative testing results during March, April, May and June combined (a), as well as separately in March (b), April (c), May (d) and June (e). Blue and red dots represent positive and negative RT-PCR results, respectively. The black circle depicts the high desity positive RT-PCR region. The singleton cluster on the right of the UMAP embeddings includes 105 patients with 90% feature values missing in their profile vectors. Those missing values are imputed as the overall mean of each feature, which makes those profiles almost identifcal to each other. Since UMAP preserves the pairwise similiarity during the mapping process, these vectors are mapped to a tiny crowd, which was excluded from our next analysis. Percentage of positive RT-PCR within and outside the circle, and percentage of negative RT-PCR within and outside the circle, are shown in the table, respectively.

To characterize the COVID19-TLP, we investigated the distribution of each laboratory test corresponding to the positive and negative RT-PCR results within and outside the circle, respectively. Violin plots of representative laboratory tests (**Supplemental Figure 2**) show, for example that COVID-19 patients presenting in the ED, as part of the COVID19-TLP, had lower absolute lymphocyte, monocyte and basophil counts, hypocalcemia, and higher red blood cell counts as well as higher hemoglobin levels and hematocrits compared to the SARS-CoV-2 negative ED patients. While no single laboratory test can accurately discriminate SARS-CoV-2 infected from uninfected patients, the combination of 21 laboratory tests formed a distinct profile that characterized typical SARS-CoV-2 positive ED patients, separating them from the SARS-CoV-2 negative ED patients.

As shown in **Figure 3**, overall the C_*T*_ values of SARS-CoV-2 RT-PCR results demonstrated an increasing trend (i.e. decreasing viral load) from April to June (C_*T*_ values in March were excluded from the analysis as they were generated from the Altona REALStar instrument with values that were not directly comparable with the other RT-PCR instruments (21)). The RT-PCR results within the circle had lower C_*T*_ values compared to those outside the circle (mean ± SD: 28.3±5.0 vs. 32.4±7.6, median: 28.7 vs 33.0, p < 0.001). In other words, higher viral loads were seen in SARS-CoV-2 positive patients who had the COVID19-TLP compared to other positive patients who did not.

**Figure 3.**
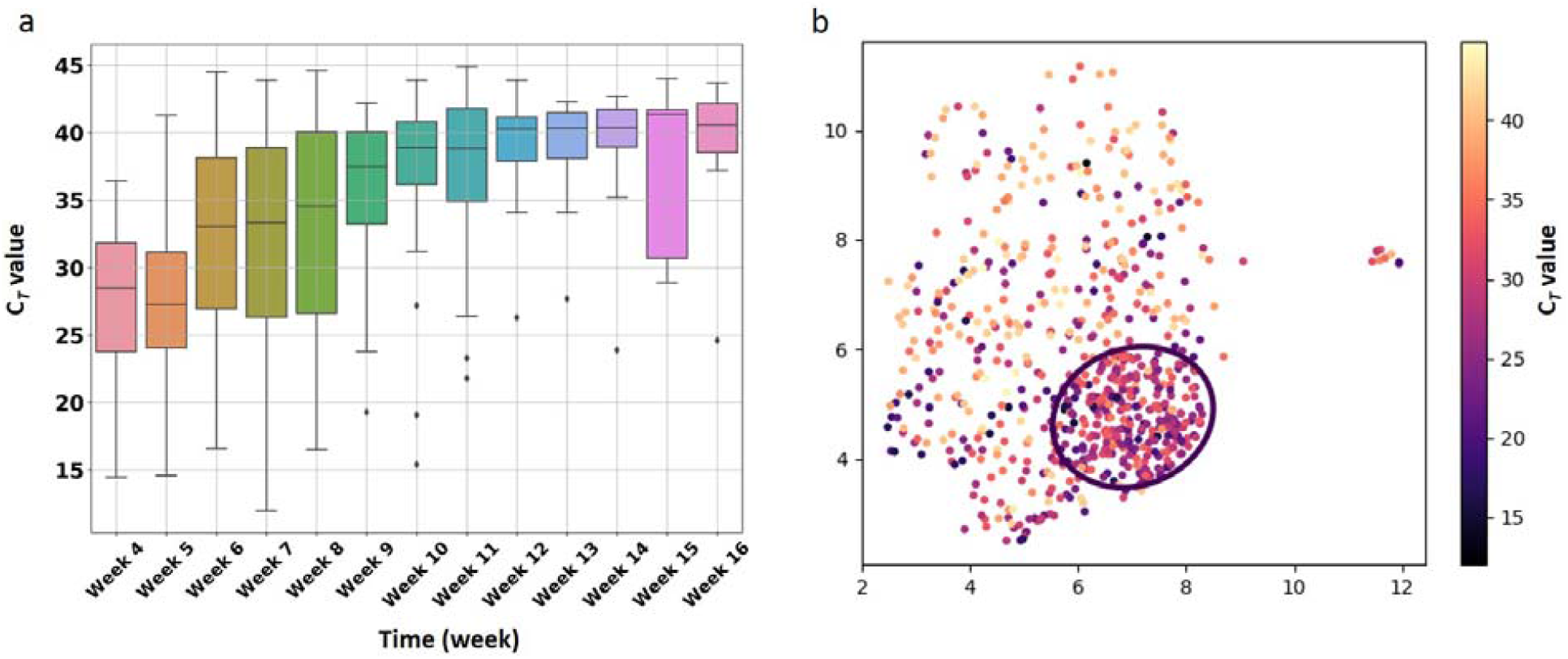
Trend of the SARS-CoV-2 RT-PCR cycle threshold (C_*T*_) values for the SARS-CoV-2 specific target. (a) Box plot the C_*T*_ values in each week from April to June. (b) UMAP analysis of the C_*T*_ value associated with the SARS-CoV-2 RT-PCR results. The black circle is the same as in Figure 2. Color bar shows the SARS-CoV-2 RT-PCR C_*T*_ value from low (black) to high (yellow).

Chart reviews were performed to investigate the clinical outcome of each SARS-CoV-2 positive patient, including whether they were discharged from the ED or admitted to an inpatient ward, whether they required care in the ICU, whether they developed respiratory failure and were intubated, and whether they died or survived COVID-19. Twenty-one patients who were transferred to other hospitals were excluded due to unknown outcomes. Overall, SARS-CoV-2 positive patients with the COVID19-TLP in our dataset had a higher incidence of hospital admission (95.7% vs. 78.4%, p < 0.001), ICU admission (27.2% vs. 15.2%, P < 0.001) and intubation (24.7% vs. 11.5%, p < 0.001) than SARS-CoV-2 patients without the COVID19-TLP, where the p-values were obtained after age adjustment. For the patients who had been admitted, the length of stay in hospital was significant longer in SARS-CoV-2 patients with the TLP than the other positive patients without the TLP (mean ± SD: 16.6±22.1 vs. 12.7 ±21.0, median 8 vs. 5 days, p < 0.001).

We further investigated the patients who had negative RT-PCR results, but had laboratory testing results that mapped within the circle (n = 116). Among them, 48 patients presented to the ED with COVID-like symptoms such as fever, cough, dyspnea and/or malaise, and 3 were reported to have close contacts with persons who tested positive for SARS-CoV-2. Nine patients (7.8% of the 116 patients) were diagnosed with COVID-19 within two days upon repeated RT-PCR testing (majority of patients tested negative did not have a repeated testing) and four other patients (2.5%) tested positive for COVID-19 antibodies one to two months after their ED visit. Therefore, the combination of specific laboratory testing results may identify some SARS-CoV-2 infected patients with a false negative RT-PCR result. Three patients were diagnosed with another respiratory virus infection such as influenza A or human rhinovirus/enterovirus.

## Discussion

In this study, using machine learning analysis, we show that approximately half of the SARS-CoV-2 positive ED patients had a distinct profile of routine laboratory test results that clearly separate them from the SARS-CoV-2 negative patients. Notably, the SARS-CoV-2 patients with the COVID19-TLP had an overall higher viral load and poorer clinical outcome compared to the other positive patients without the COVID19-TLP. The identification of COVID-19 distinct laboratory profile could be used to prioritize high-risk patients, assisting in ED patient triaging and optimizing the usage of resources in areas where RT-PCR testing is not accessible due to financial or supply constraints. Furthermore, our temporal analysis illustrates the substantial decrease in the percentage of patients with the COVID19-TLP in May and June 2020, after the initial surge of COVID-19 in March and April 2020, in NYC. The observed trend in the laboratory result profile provides insight to the epidemiologic and biologic evolution of the disease, which could play an important role in COVID-19 population disease severity tracking and prediction and may assist in directing public health policies as COVID-19 spreads to new geographic areas or as a “second wave” occurs in previously affected areas.

Existing research has shown that the SARS-CoV-2 viral load correlates with severity of COVID-19 presentation (22), and is independently associated with an increased risk of intubation and/or in-hospital mortality (23), (24, 25). Here, we demonstrate that SARS-CoV-2 viral load also correlates with a panel of laboratory test result abnormalities (COVID19-TLP). Patients who have a higher viral load and a COVID19-TLP at presentation may have a higher risk of adverse outcomes. Thus, our analysis provides a means of identifying patients with more severe physiologic disturbance and poorer outcome. Analysis of the laboratory profile at ED presentation provides complementary information, which, because of the rapid turn-around-time (usually within a couple of hours) for routine laboratory test results, offers an opportunity for rapid triaging and more timely intensive monitoring of high-risk patients. In addition, this analysis may also suggest which patients are unlikely to be SARS-CoV-2 positive, as overall 97% of SARS-CoV-2 negative patients were outside the circle (did not have the COVID19-TLP). As such, this analysis could be deployed clinically as an application integrated into the electronic medical record (EMR) system and visually show if the dot corresponding to an individual patient is within or outside the circle as soon as the patient’s laboratory test results are available. In areas where SARS-CoV-2 RT-PCR is not accessible onsite, this analysis may provide a timely clue to prioritize high-risk patients.

Laboratory tests provide an objective and quantifiable means to characterize the evolution of COVID-19. In addition to an overall decrease in the number of positive cases, our study depicts a declining trend in the viral load of SARS-CoV-2 patients as well as a decreasing percentage of patients showing the COVID19-TLP from April to June 2020. In our hospital, RT-PCR tests in March and April were primarily offered to symptomatic patients due to a limited testing capacity. Testing was expanded to more patients, both symptomatic and asymptomatic, in May and June when supplies, equipment and testing personnel were available. While more widely available testing in May and June may contribute to the decrease in the percentage of severe patients, it is unclear whether there are other contributing factors such as changes in virus virulence, modifications of population behavior by adhearing to public health directives such as wearing masks, increased patient awareness of the disease with physician visits sooner after symptom onset (presumably associated with lower viral loads), a decrease in the number of most vulnerable patients as they have already been infected. Our analysis, based upon a patient population in NYC during the peak of COVID-19, provides to researchers, physicians and public health authorities an insightful method to better understand the evolution of this disease from a laboratory testing perspective. In addition, our model based on laboratory test results reflecting the physiologic effects of the virus on patients, may improve our understanding of the pathobiology of the SARS-CoV-2, and thus, aid in devising guidances for treatment, tracking and prevention of COVID-19.

Our study has a limitation that the analysis of patient data was performed at a single large metropolitan medical center. Therefore, the role of the COVID19-TLP in discriminating between SARS-CoV-2 negative and positive patients should be tested on a larger scale at other medical centers in areas with varying degrees of COVID-19 prevalence.

## Conclusions

Using machine learning analysis, we have identified a typical laboratory test result profile for SARS-CoV-2 positive patients, which correlates with higher viral load and poorer clinical outcome. Overall 97% of the SARS-CoV-2 negative patients did not have the COVID19-TLP. This analysis could serve as an important tool to prioritize high-risk patients and optimize the usage of resource. Furthermore, this analysis illustrates the down-trending in the proportion of SARS-CoV-2 patients with the COVID19-TLP from the initial surge of COVID-19 to a later post-apex phase in NYC, the intial epicenter of the pandemic in the US. Our findings have shed new light on the evolution and pathobiology of COVID-19.

## Data Availability

Data in the manuscript are available upon request.

## Abbreviation

COVID-19: COVID-19
COVID19-TLP: COVID19-TLP
C_*T*_: C_*T*_
ED: ED
ICU: ICU
MCV: MCV
NYPH/WCMC: NYPH/WCMC
RDW-CV: RDW-CV
RT-PCR: RT-PCR
SARS-CoV-2: SARS-CoV-2
TAT: TAT
UMAP: UMAP
WBC: WBC

## Acknowledgement

We want to thank Hanna Rennert and Arryn R. Craney for their effort on RT-PCR method development, and medical technologists at NYPH/WCMC who performed the laboratory testing.

## Author contribution

HSY for conceptualization, investigation, data collection and analysis, writing the original draft and editing of the manuscript. YH and HZ for data analysis, visualization and editing the manuscript. AC and LFW for conceptualization and editing the manuscript. RF for organzing the dataset and performing data analysis. SRB for editing the manuscript. PV for providing RT-PCR data. JLS for providing C_*T*_ values and editing the manuscript. MMC, RK and ZZ for reviewing and editing the manuscript. FW for conceptualization, supervision of the project, investigation, data analysis, and editing the manuscript.

---

**Supplementary Figure 1.**
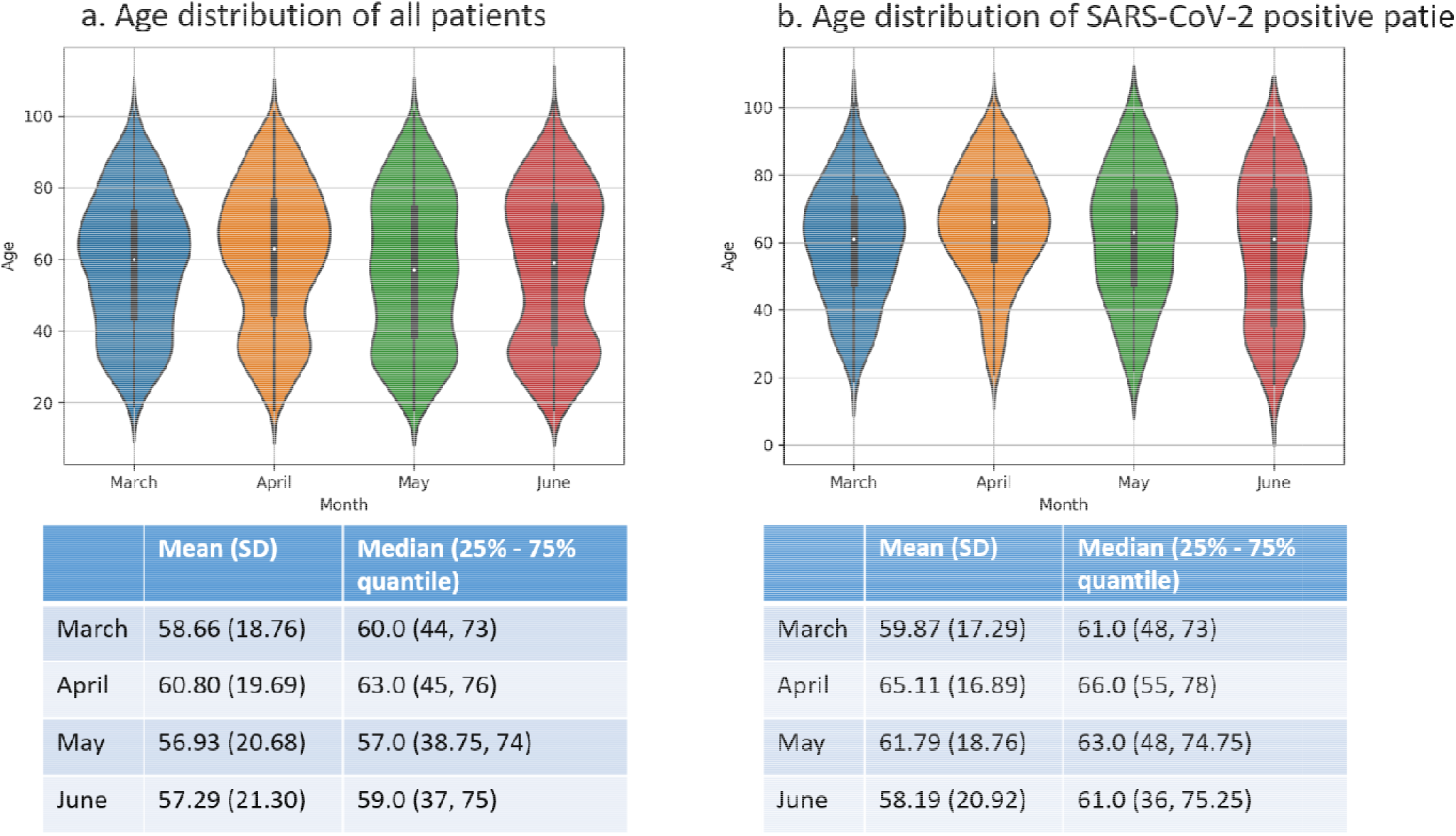
Distribution of age in total RT-PCR tested patients (a) and SARS-CoV-2 positive patients (b) in March, April, May and June. Mean (SD) and median (25% - 75% quantile) are shown under each figure.

**Supplementary Figure 2.**
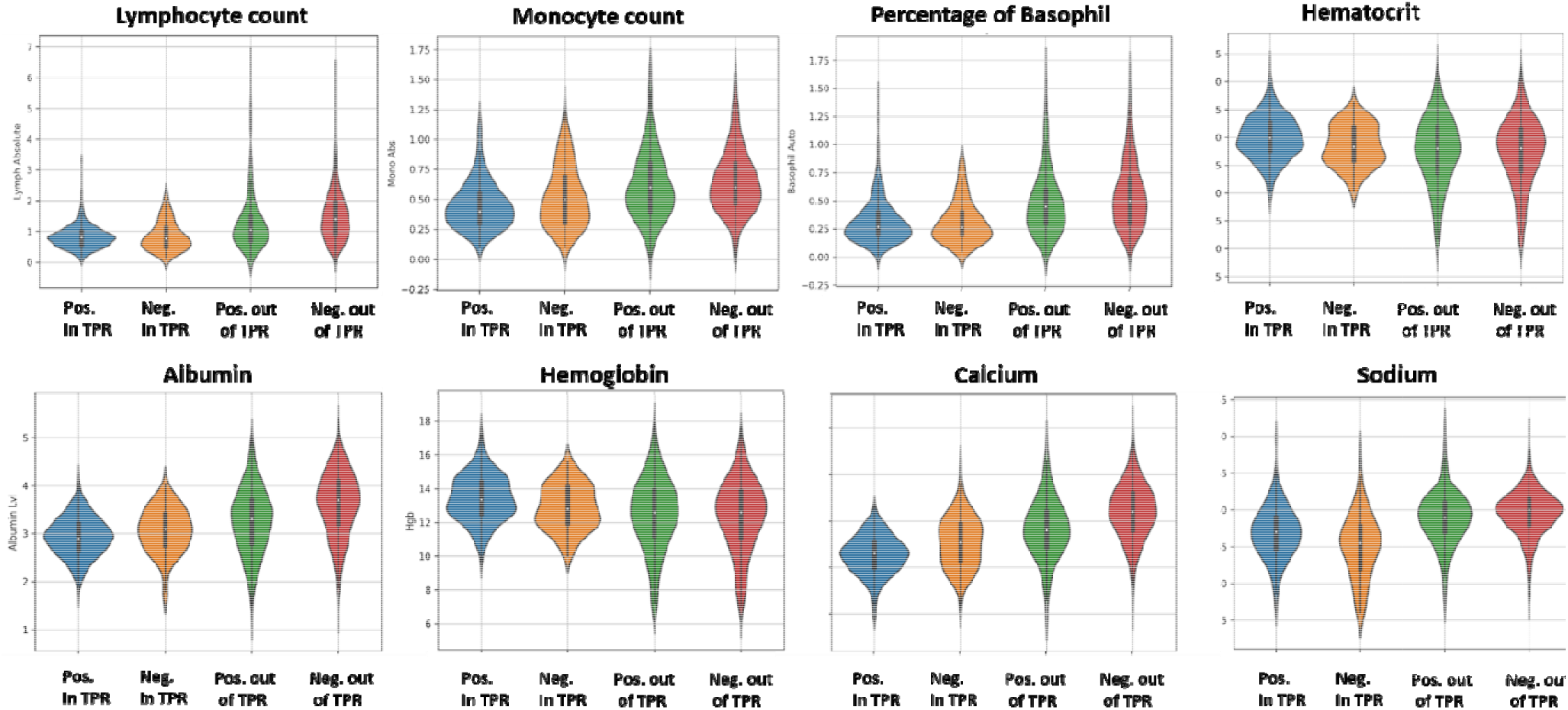
Distribution of representative laboratory tests of positive RT-PCR within the TPR, negative RT-PCR within the TPR, positive RT-PCR outside the TPR, and negative RT-PCR outside the TPR.

